# Health impact assessment of ambient fine particulate matter 2.5 (PM_2.5_) decline during COVID-19 movement restrictions, India

**DOI:** 10.64898/2025.12.31.25343296

**Authors:** Ankita S Achanta, Ther W Aung

**Affiliations:** Department of Nutrition, School of Medicine, Case Western Reserve University, 10900 Euclid Ave, Cleveland, Ohio, 44106, United States of America (USA); The MetroHealth System, Case Western Reserve University, Cleveland, USA

## Abstract

**Objective:** To quantify avoidable deaths in five Indian megacities associated with reductions in ambient levels of particulate matter 2.5 µm or less in diameter (PM_2.5_) during two coronavirus disease 2019 (COVID-19) movement restrictions.

**Methods:** We obtained data from United States’ Embassy and Consulate monitors on PM_2.5_ concentrations in Chennai, Hyderabad, Kolkata, Mumbai and New Delhi during the first (25 March to 31 May 2020) and second (5 April to 15 June 2021) COVID-19 movement restrictions. We used the World Health Organization’s AirQ+ tool to estimate the annual number of PM_2.5_-associated avoidable deaths over the long term due to all causes, chronic obstructive pulmonary disease, lung cancer, ischaemic heart disease and stroke in adults and due to acute lower respiratory infection in young children.

**Findings:** Mean PM_2.5_ concentrations across the five cities decreased by 28– 77% during the first movement restriction and by 48–69% during the second movement restriction. During the first movement restriction alone, decreases in PM_2.5_ were associated with an estimated 132□000 fewer annual deaths from all causes across the five cities and with 11□100, 3900, 5500, 3900 and 1240 fewer annual deaths due to chronic obstructive pulmonary disease, lung cancer, ischaemic heart disease, stroke and acute lower respiratory infection, respectively.

**Conclusion:** Improvements in air quality equivalent to those observed during COVID-19 movement restrictions in Indian megacities could save hundreds of thousands of lives annually. Higher air quality standards and aggressive mitigation and enforcement measures are needed to reduce mortality due to air pollution.

## Introduction

India has one of the highest public health burdens from air pollution.^1,2^ An estimated 1.7 million deaths (18% of all deaths in the country) were attributable to ambient and household air pollution in 2019, with the former accounting for most of these deaths.^3^ The health burden of air pollution is associated with substantial economic costs for the country: economic losses due to premature death and morbidity in 2019 were estimated to be 29 billion United States dollars (US$) and US$8 billion, respectively.^4^

In 2019, the Indian government launched the National Clean Air Programme to prevent, control and reduce air pollution in cities exceeding national ambient air quality standards by increasing monitoring and banning the use of high-pollution fuels.^5^ Recently, the government revised its target for the reduction in particulate matter, a key determinant of the effect of air pollution on health; the aim was to reduce the level by 40% by 2026 relative to 2017.^6^ However, progress on meeting these air pollution goals has been inadequate and they are challenging, particularly in urban environments; recent reports note that the decline in air pollution from 2017 to 2021 has been lower than expected, with some cities experiencing an increase.^7,8^

Urban air quality can be affected by a variety of local and non-local emission sources and by meteorological conditions.^9^ Several source apportionment studies in Indian cities have shown that local sources, such as resuspended dust (e.g. due to wind or surface abrasion), vehicular emissions, secondary aerosols, fossil fuel combustion and residential biomass use, contribute most to urban ambient particulate pollution.^10^ A systematic review of local source apportionment studies in India found that traffic was the largest contributor to urban pollution with ambient particulate matter 2.5 µm or less in diameter (PM_2.5_), accounting for 37%.^11^ Non-local sources included agricultural biomass burning, particularly around New Delhi.^12^

The importance of local sources of particulate matter for urban ambient air quality became evident during the coronavirus disease 2019 (COVID-19) movement restrictions when overall anthropogenic activity, including regular automobile use, decreased dramatically and many cities reported substantial improvements in air quality.^13,14^ In India, several studies based on a variety of air monitoring data sources, including local monitoring networks and satellite data, reported better urban air quality during movement restrictions.^15–17^ However, these studies were limited to an individual city or movement restriction period, which made spatial and temporal comparisons difficult.

Further, the quantitative health impact of improvements in air quality during movement restrictions in India has not previously been reported. Both acute and chronic exposure to PM_2.5_ is associated with premature death, birth defects, lung cancer and the development of respiratory, metabolic, neurological and cardiovascular diseases.^18^ Children are especially sensitive to these particle because of their lower body mass relative to adults, with children younger than 5 years being especially susceptible to death from acute lower respiratory infection.^19^

The aims of our study were to quantify the reduction in ambient PM_2.5_ concentrations that occurred during the two COVID-19 movement restriction periods in the five major Indian cities and to estimate the long-term health benefits attributable to the resulting improvements in air quality. We hope our findings will assist policy-makers in evaluating the cost of different approaches to mitigating particulate matter pollution, given the potential health benefits for millions of India’s urban residents.

## Methods

The study involved five geographically dispersed, densely populated, metropolitan areas in India: Chennai, Hyderabad, Kolkata, Mumbai and New Delhi. These five cities, which were selected because of their recognized poor air quality, had over 88 million residents in 2021.^18,20^ In 2020 and 2021, the Indian government implemented two periods of movement restrictions across these five cities to control the spread of severe acute respiratory syndrome coronavirus 2 (SARS-CoV-2). The first period took place from 25 March 2020 to 31 May 2020 (duration: 68 days) and the second, from 5 April 2021 to 15 June 2021 (duration: 72 days). The movement restriction mandate required all citizens to stay at home, except those working in essential services. This approach resulted in a near complete shutdown of normal anthropogenic activities. For comparison, we considered duration-matched control periods before and after the first and second movement restriction period (Table 1).

**Table 1.** Study time periods, health impact of changes in ambient PM_2.5_ concentrations during COVID-19 movement restrictions, India, 2020–2021.

| Study period | Date range | Duration of study period, days |
| --- | --- | --- |
| Before first movement restriction | 17 January to 24 March 2020 | 68 |
| During first movement restriction | 25 March to 31 May 2020 | 68 |
| After first movement restriction | 1 June to 7 August 2020 | 68 |
| Before second movement restriction | 23 January to 4 April 2021 | 72 |
| During second movement restriction | 5 April to 15 June 2021 | 72 |
| After second movement restriction | 16 June to 26 August 2021 | 72 |

COVID-19: coronavirus disease 2019; PM<sub>2.5</sub>: particulate matter $\leq 2.5$ $\mu\text{m}$ in diameter.

### PM_2.5_ concentrations

Two main ground-based sources of PM_2.5_ data for air quality research exist in India: (i) the Indian Central Pollution Control Board; and (ii) the embassies and consulates of the United States of America. For our study, we used United States’ Embassy and Consulate data rather than Central Pollution Control Board data, which had problems with reliability and inconsistent data reporting.^21^ These embassies and consulates used standardized, high-quality instruments approved as a United States’ Federal Equivalent Method: Met One Beta Attenuation Mass Monitors (Met One Instruments, Grants Pass, USA).^21^ The data are publicly available from AirNow.^22^ For each study city, we downloaded hourly raw data on PM_2.5_ concentrations for the years 2017 to 2021. The data were cleaned and processed for analysis using the open-source pollucheck v. 1.0 package,^23^ which is an application for analysing and visualizing air quality data.

### Health impact analysis

To model the number of avoidable annual deaths resulting from a reduction in particulate matter pollution, we used updated World Health Organization (WHO) air quality guidelines for PM_2.5,_ which were released in 2021,^24^ and a recently revised version (v. 2.2) of WHO’s AirQ+ tool,^25^ which incorporates a health impact function for estimating health outcomes attributable to improved air quality. The health impact function provides an estimate of the annual number of avoidable premature illnesses or deaths that could result from an improvement in air quality in a defined geographical location and is expressed as:

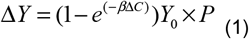

where Δ*Y* is the estimated number of avoidable premature illnesses or deaths; β is□a risk estimate (or β coefficient) obtained from epidemiological studies; Δ*C* is the change in the concentration of the air pollutant being examined; *Y*_*0*_ is the baseline rate (i.e. incidence) of illnesses or deaths; and *P* is the size of the population exposed to air pollution. Information required for the AirQ+ analysis includes: (i) air pollutant concentrations obtained by monitoring; (ii) population data; (iii) a baseline disease or death rate; and (iv) a concentration–response parameter, such as a β coefficient, that quantifies the likelihood of a health effect associated with a change in the concentration of the air pollutant.

We limited our analysis to the long-term impact on health because implementation of air pollution and emission control policies are likely to extend beyond the duration of the COVID-19 movement restrictions. Moreover, a long-term impact assessment is useful for planning emission control measures and for evaluating the costs and benefits of different mitigation strategies.

We analysed all-cause mortality in individuals of both sexes aged 25 years and older and five specific causes of death: (i) chronic obstructive pulmonary disease in individuals aged 25 years and older; (ii) lung cancer in individuals aged 25 years and older; (iii) ischaemic heart disease in individuals aged 50 to 74 years; (iv) stroke in individuals aged 50 to 74 years; and (v) acute lower respiratory infection in children younger than 5 years. The age range of 50 to 74 years was chosen for ischaemic heart disease and stroke after considering the prevalence and incidence of these conditions in the whole Indian population.

In the AirQ+ tool, the standard relative risk for the concentration–response function for all-cause mortality is 1.08 (95% confidence interval, CI: 1.06–1.09) for an increase in annual PM_2.5_ concentration of 10 μg/m^3^. For each defined time period, we estimated the health impact of an observed mean daily PM_2.5_ concentration by comparison with a counterfactual, or reference, PM_2.5_ concentration of 5 μg/m^3^, which is the maximum annual value recommended by the 2021 WHO air quality guidelines.^24^

We obtained population data for all five cities for 2020 to 2021 from the World Population Review website.^20^ For baseline rates of relevant health outcomes by age, sex and year, we used data from the 2019 Indian Global Burden of Disease study.^26^ Details of the population age distribution, the relative risk and incidence of different conditions, and other data required for the AirQ+ analysis are available from the online repository.^27^

## Results

### Daily PM_2.5_ concentrations

Before the first movement restriction period, Chennai recorded the lowest mean daily PM_2.5_ concentration (28 μg/m^3^), while Kolkata had the highest (110 μg/m^3^; Table 2). During the movement restriction periods, the mean daily concentration decreased markedly in all five cities, with Kolkata and Mumbai experiencing the largest falls. The reduction in mean PM_2.5_ concentration from before to during the first movement restriction ranged from 28% (from 43 μg/m^3^ to 31 μg/m^3^) in Hyderabad to 77% (from 110 μg/m^3^ to 25 μg/m^3^) in Kolkata; from before to during the second movement restriction, the reduction ranged from 48% % (from 121 μg/m^3^ to 63 μg/m^3^) in New Delhi to 69% % (from 118 μg/m^3^ to 36 μg/m^3^) in Kolkata. The PM_2.5_ concentrations measured after the movement restriction periods indicate that either the reductions were maintained or that concentrations stayed below pre-movement restriction levels.

**Table 2.** Mean daily PM_2.5_ concentrations in study cities, health impact of changes in ambient PM_2.5_ concentrations during COVID-19 movement restrictions, India, 2020–2021.

| City | Time period<sup>a</sup> |  |  |  |  |
| --- | --- | --- | --- | --- | --- |
|  | Before first movement restriction | During first movement restriction |  | After first movement restriction | Before second movement restriction |
| | Mean daily PM<sub>2.5</sub> concentration, $\mu\text{g}/\text{m}^3$ (95% CI) | Mean daily PM<sub>2.5</sub> concentration, $\mu\text{g}/\text{m}^3$ (95% CI) | Change from before first movement restriction, % | Mean daily PM<sub>2.5</sub> concentration, $\mu\text{g}/\text{m}^3$ (95% CI) | Mean daily PM<sub>2.5</sub> concentration, $\mu\text{g}/\text{m}^3$ (95% CI) |
| Chennai | 28 (26–31) | 14 (12–16) | –50 | 16 (15–17) | 37 (34–41) |
| Hyderabad | 43 (38–47) | 31 (29–33) | –28 | 13 (11–14) | 70 (65–74) |
| Kolkata | 110 (98–122) | 25 (22–28) | –77 | 14 (12–16) | 118 (106–130) |
| Mumbai | 70 (63–77) | 24 (22–27) | –65 | 15 (14–17) | 66 (61–71) |
| New Delhi | 101 (89–112) | 44 (40–48) | –56 | 45 (41–49) | 121 (108–134) |

CI: confidence interval; COVID-19: coronavirus disease 2019; PM<sub>2.5</sub>: particulate matter $\leq 2.5$ μm in diameter.
<sup>a</sup> Details of the time periods are shown in Table 1.

Fig. 1 illustrates the percentage of days before, during and after the two movement restrictions when the PM_2.5_ concentration was in different health impact categories, as classified by the Indian Government’s Air Quality Index.^28^ The Air Quality Index defines six air quality categories ranging from good to severe in terms of health impact. Good air quality was defined as a PM_2.5_ concentration below 50 μg/m^3^ and severe air quality was defined as a concentration between 401 and 500 μg/m^3^. Generally, the percentage of days when the air quality was good or satisfactory (i.e. a PM_2.5_ concentration below 100 μg/m^3^) in the five cities was higher during movement restrictions and post-movement restriction periods than during pre-movement restriction periods. Moreover, the percentage of days that met Air Quality Index daily PM_2.5_ concentration standards was markedly higher after each movement restriction period in all cities. In particular, Kolkata had relatively bad air quality before the first and second movement restrictions, as indicated by the percentage of days classified as having moderate or poor air quality (i.e. PM_2.5_ concentration: 101 to 300 μg/m^3^). After each movement restriction period, air quality in Kolkata was classified as good or satisfactory (i.e. PM_2.5_ concentration below 100 μg/m^3^) on all days.

We also compared mean 24-hour PM_2.5_ concentrations in the five cities with the 24-hour concentrations specified in WHO’s air quality guidelines to progressively guide efforts to meet these guidelines.^24,28^ According to WHO’s guidelines, the 24-hour PM_2.5_ concentration should not exceed 15 µg/m^3^. In addition, there are four interim 24-hour targets: (i) interim target 1 is 75 μg/m^3^; (ii) interim target 2 is 50 μg/m^3^; (iii) interim target 3 is 37.5 μg/m^3^; and (iv) interim target 4 is 25 μg/m^3^. Before the movement restriction periods, PM_2.5_ concentrations in the cities were generally six to eight times higher than recommended by WHO’s air quality guidelines. However, Chennai, Hyderabad and Mumbai had already met some interim targets. The movement restrictions brought all cities within range of the interim targets and closer to the PM_2.5_ concentration recommended by the air quality guidelines. For example, all cities, except New Delhi, met the more stringent, interim targets 3 and 4. In New Delhi, despite an approximately 50% reduction in the mean daily PM_2.5_ concentration during the movement restriction periods, PM_2.5_ levels met only interim targets 1 and 2.

Improvements in air quality were seen even in the post-movement restriction periods after the movement restriction mandates had ended. After the first movement restriction period, all cities had good or satisfactory air quality, as classified by the Indian National Ambient Air Quality Standard.^29^ Except for New Delhi during the second movement restriction period, all cities met or exceeded the 24-hour Indian National Ambient Air Quality Standard for PM_2.5_ of 60 μg/m^3^ during the two movement restriction periods.^29^

### Health impact

In New Delhi, which had some of the highest PM_2.5_ concentrations during the study period, we estimated that before the second movement restriction period, PM_2.5_ was associated with 131□405 (95% CI: 109□326–140□652) avoidable deaths per year from all causes (Table 3). Among the specific causes of death, chronic obstructive pulmonary disease was associated with the highest number of avoidable deaths per year across all cities, followed by ischaemic heart disease.

**Table 3.**
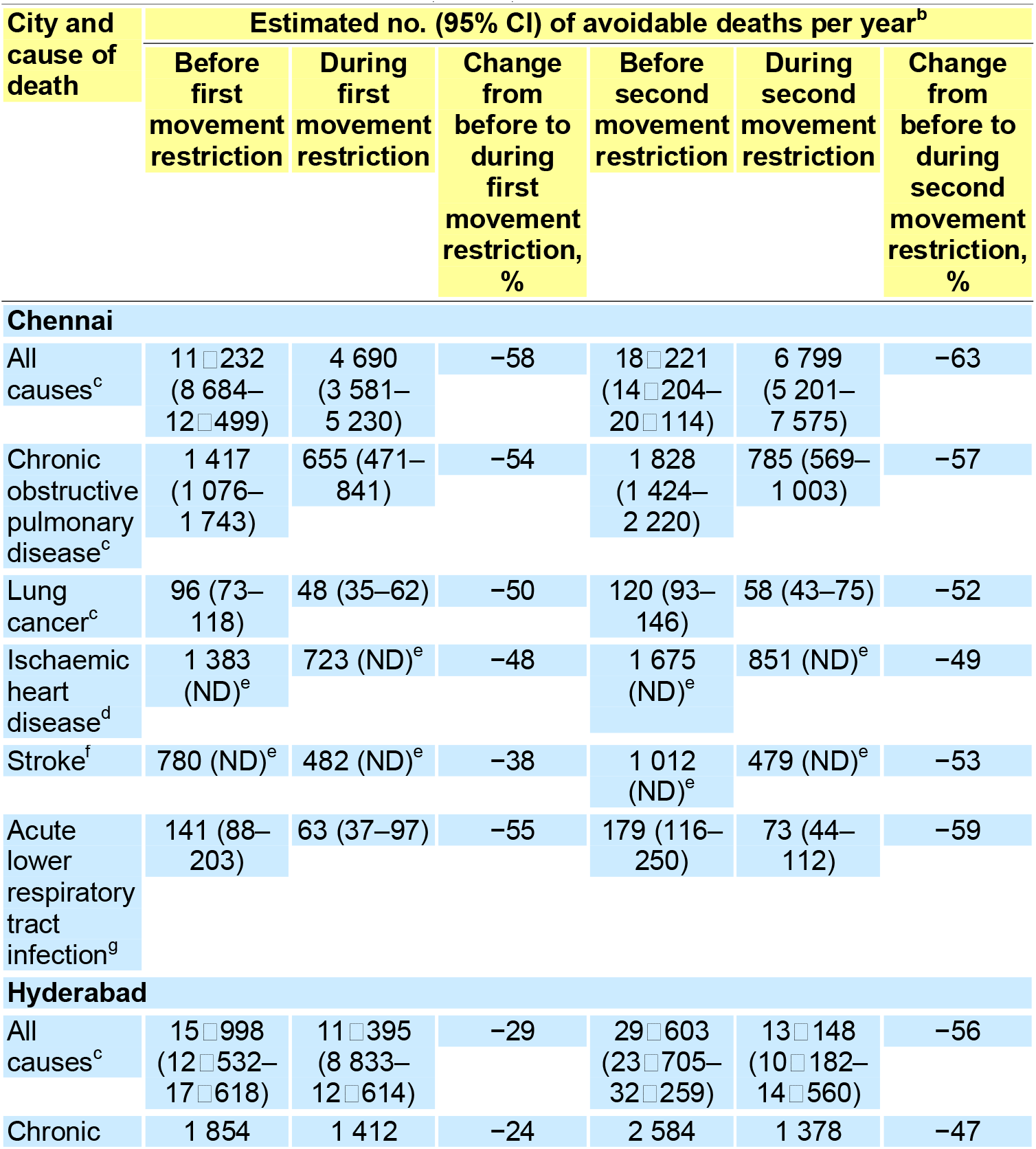
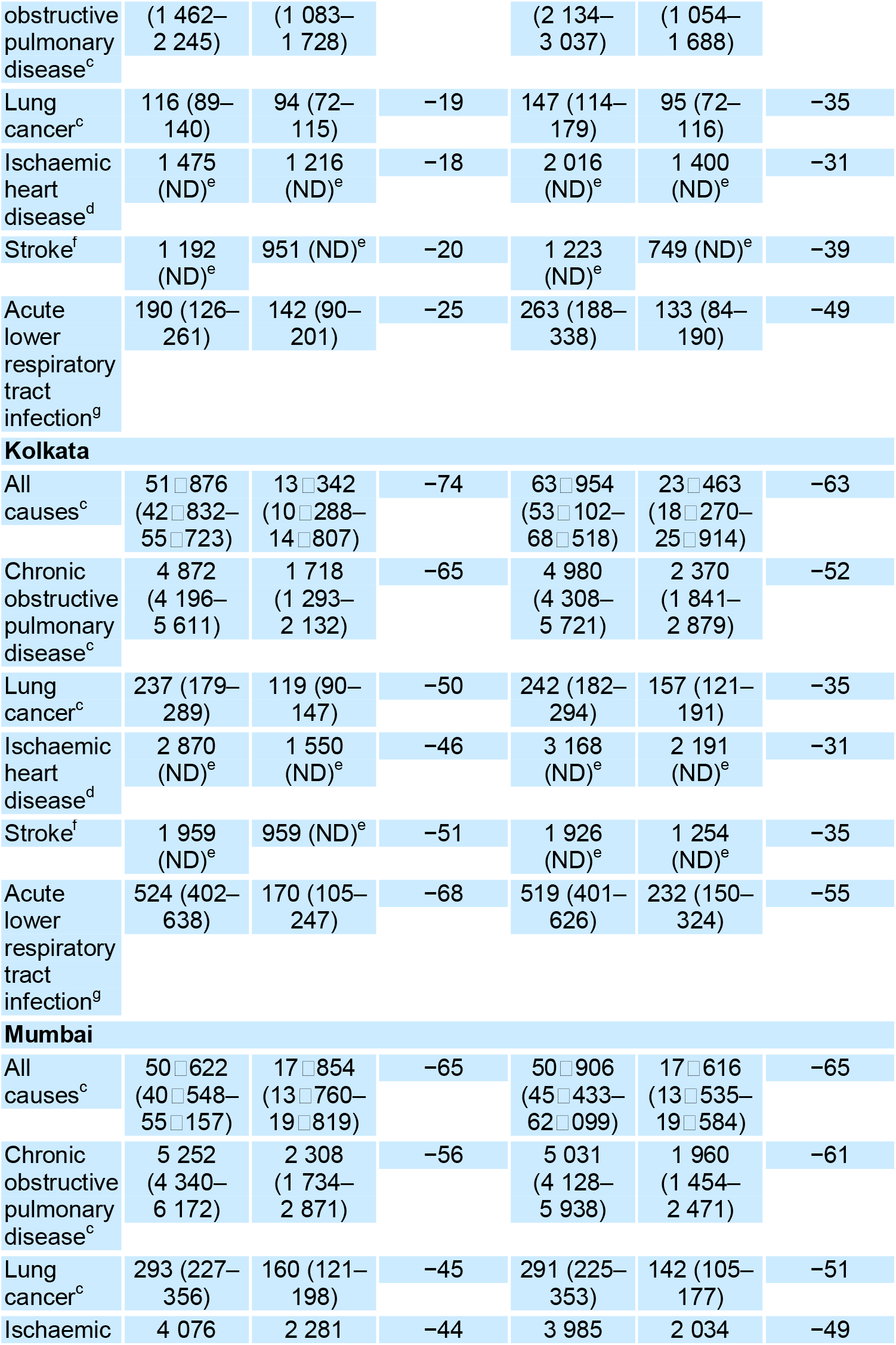
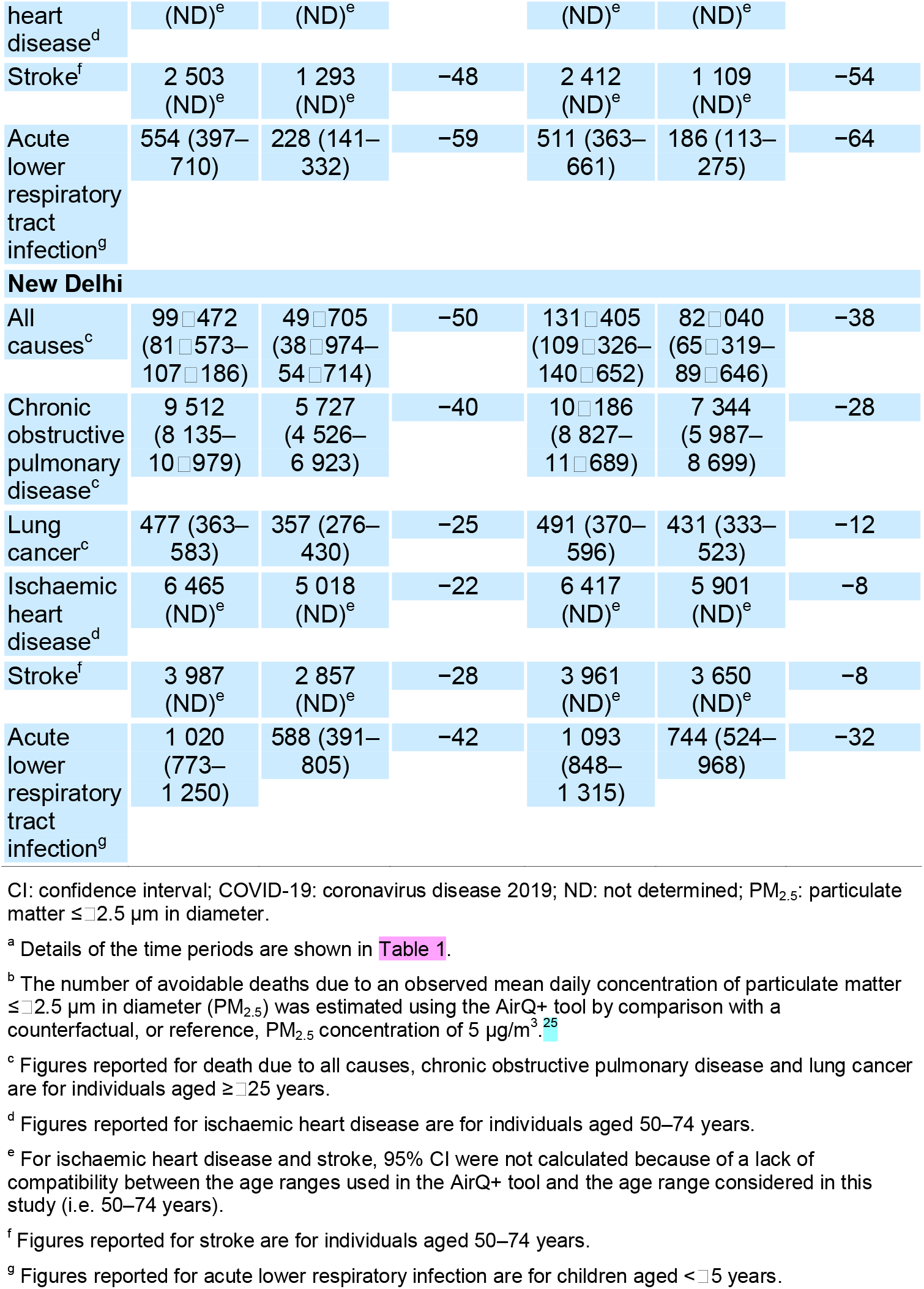
Avoidable deaths due to PM_2.5_ pollution, by city, cause of death and time period,^a^ health impact of changes in ambient PM_2.5_ concentrations during COVID-19 movement restrictions, India, 2020–2021.

During both movement restriction periods, the reductions in PM_2.5_ concentrations observed were associated with a lower number of deaths for all health outcomes analysed across the five cities. We estimated that, during the first restriction period, the number of deaths due to all causes decreased by approximately 132□000 per year across all cities. For specific causes of death, the corresponding annual reductions were estimated to be approximately 11□100 for chronic obstructive pulmonary disease, 3900 for lung cancer, 5500 for ischaemic heart disease, 3900 for stroke and 1240 for acute lower respiratory infection. The decline in all-cause mortality during the first movement restriction ranged from 29% (from 15□998 to 11□395 deaths) in Hyderabad to 74% (from 51□876 to 13□342 deaths) in Kolkata. The large decline in the estimated number of deaths for Kolkata is unsurprising given that the city had the highest mean daily PM_2.5_ concentrations before the first movement restriction period and the greatest reduction in concentrations during the first restriction period. However, some reductions in the estimated number of avoidable deaths were minimal. For example, estimates for New Delhi suggest that there was only an 8% decline in the number of deaths due to ischaemic heart disease and stroke (from 6417 to 5901 deaths and from 3961 to 3650 deaths, respectively) during the second movement restriction.

## Discussion

The COVID-19 movement restrictions in 2020 and 2021 considerably lowered PM_2.5_ pollution, to a varying extent, in the five Indian megacities we investigated. Variations may have been due to each city’s unique social, political and geographical characteristics, meteorological conditions and dominant sources of pollution. For example, in New Delhi, there were days with only moderate air quality during the second movement restriction, which may have been due to non-traffic emission sources.

Our findings are in line with other studies that reported improvements in air quality during COVID-19 movement restrictions. One previous study in India, which used data from the United States’ Environmental Protection Agency for only the first movement restriction, documented changes in air quality in the same five cities we studied.^30^ That study’s authors found that mean declines in PM_2.5_ concentrations during the first movement restriction ranged from 10 to 62%, slightly lower than in our analysis (i.e. 28 to 77%). We attribute the difference to different definitions of the period before the first movement restriction: a period of 24 days was used in the previous study compared with 68 days in our analysis.

Nevertheless, we believe both approaches provide a valid illustration of the effect of movement restrictions on ambient PM_2.5_ concentrations. The mean PM_2.5_ concentrations reported during the first movement restriction period in the previous study were nearly identical to the levels we found in all five cities.

Seasonal effects may also have contributed to the changes in air pollution we observed because the periods before, during and after the two movement restrictions covered January to August and the transition from winter to the monsoon season. Although PM_2.5_ concentrations vary by location in India, the highest levels generally occur during winter, after which they decline in spring and summer and reach their lowest during the monsoon.^31^ However, a comparison of PM_2.5_ concentrations from 2017 to 2019 (before the COVID-19 pandemic) in the months corresponding to the movement restriction months in 2020 and 2021 indicated that the movement restrictions were associated with additional reductions in PM_2.5_ (details are available from the online repository).^27^

We use the scenario of reduced PM2.5 reductions that occurred during the COVID-19 movement restrictions to model long-term health benefits achievable in India’s major cities. However, our findings probably underestimated the true total health benefits because PM_2.5_ has additional adverse health impacts that are currently not included in the AirQ+ tool and have not been quantified here, such as impacts on birth outcomes, diabetes, cognitive decline and mental health.^3,4^

The study has limitations. First, the United States’ Embassy and Consulate air quality monitors were generally located in affluent areas of the study cities, where the air was relatively clean compared with lower-income areas or industrial zones. Moreover, the PM_2.5_ concentrations used in our study were obtained using only one monitor in each city (in the United States’ facility). Consequently, actual particulate matter levels across the megacities were likely to have been underestimated. Second, epidemiological studies specific to India on the effect of increased PM_2.5_ concentrations on mortality are lacking. The relative risk estimates used by the AirQ+ tool come from studies conducted in high-income countries. Consequently, locally relevant, epidemiological studies in low- and middle-income countries are needed.

Nevertheless, our study demonstrates that the AirQ+ tool can provide a practical means of analysing the implications of policy and, in particular, can help guide the revision of national ambient air quality standards. Moreover, as our study followed the recommendations for good reporting practice in the GATHER guidelines,^29^ our findings can be compared with data from other analyses and from different locations.^32^

Currently, the Indian government is revising its national guidelines on ambient air quality standards. To protect the population’s health, it is of importance that this process consider the health implications of air quality.^32^ For example, a recent study across 10 Indian cities indicated that PM_2.5_ concentrations considerably below 60 μg/m^3^ (the current national ambient air quality standard for the 24-hour PM_2.5_ level) were associated with substantial increases in short-term mortality.^33^ Policy-makers should consider using the interim targets specified by the 2021 WHO air quality guidelines to help achieve meaningful reductions in morbidity and mortality.^24^

A recent report on the implementation of the National Clean Air Programme in India noted that progress was unsatisfactory in 2020; one key deficiency was the absence of substantive actions to reduce emissions by capping the consumption of fossil fuels, especially in the power, industrial and transport sectors.^5^ Improvements to this programme should include enforceable mechanisms to penalize noncompliance by responsible parties and the specification of targets for reduced pollutant levels.

Our findings also highlight the need for city-specific measures. In New Delhi, efforts to improve air quality must address both vehicular emissions and seasonal agricultural burning.^32^ Recommended methods include subsidizing crop residue management technologies and promoting crop diversification.^33^ Industrial emissions are a major concern in Kolkata and Mumbai. In these cities, local strategies for improving air quality should focus on industrial clusters, with stricter penalties for noncompliance with the National Clean Air Programme.^32,33^ In Mumbai, the city's geography and vehicular congestion necessitate a robust expansion of the metro transportation network and stricter fuel standards for maritime vessels.^32^ Across all cities, road traffic emissions could be considerably reduced by: (i) expanding low-emission zones; (ii) incentivizing the adoption of electric vehicles; (iii) introducing congestion pricing in high-traffic zones; and (iv) increasing the capacity of public transit systems.^32^

Addressing sources of particulate matter is not only crucial for public health but also for mitigating climate change.^32^ WHO considers climate change as a multiplier of the adverse health impact of air pollution and has declared it to be the single biggest threat facing humanity.^3^ The Indian government has made ambitious efforts to implement climate change policies, primarily focusing on the energy sector and transitioning away from coal.^34^ Tackling local sources of urban ambient particulate matter in India’s megacities, therefore, would benefit both public health and the climate.

Our findings provide timely, real-world evidence that health should play a central role in the revision of India’s air quality standards and that tackling city-specific sources of urban PM_2.5_ emissions should be prioritized in efforts to achieve the revised National Clean Air Programme reduction targets. In addition, allocating more resources for epidemiological studies of air pollution in highly polluted and densely populated, low- and middle-income countries like India would help promote effective policy-making.

## Data Availability

All data produced in the present study are available upon reasonable request to the authors.

## Acknowledgements

We thank P Pant (Health Effects Institute, Boston, USA) and A Upadhya (ILK Laboratories, Bengaluru, India).

---

Fig. 1. **Daily air quality classifications, by city and time period, health impact of changes in ambient PM_2.5_ concentrations during COVID-19 movement restrictions, India, 2020–2021**

COVID-19: coronavirus disease 2019; PM_2.5_: particulate matter ≤□_2.5 µm in diameter.

Notes: Details of the time periods are shown in Table 1. Air quality was classified according to the Indian Government’s Air Quality Index, which defines six air quality categories ranging from good to severe in terms of health impact.^26^

